# A translational open-source flow mediated dilation assessment tool with improved automated edge detection

**DOI:** 10.1101/2025.03.18.25324231

**Authors:** Bishoy Pramanik, Daniel Veith, K.A. Fernandez, Anders J. Asp, Ryan Solinsky

## Abstract

Current methods for automated vascular edge detection are prone to motion artifact and error when there are dynamic changes in vessel diameter. This often relegates important measurement such as flow mediated dilation to the laboratory, where specialized apparatus can constrain the arm and ultrasound probe. Herein, we describe a novel open-source software program which facilitates these methods in the clinical environment with a clinical vascular ultrasonographer. We compare this software to a current, popular program for flow mediated dilation measurements, FloWave.US, and demonstrate significant improvements in retained data across a diverse study population.

---

Blood vessel diameter is an important metric in evaluating vascular function, whether it be assessing endothelial health through flow mediated dilation (FMD),^1,2^ monitoring blood vessel responses to pharmacologic stimuli,^3^ or assessing general cardiovascular status.^4^ To non-invasively collect these measurements, B-mode ultrasound footage is often dynamically recorded, with the process of measurements automated to ensure vessel wall movements from pulsatile flow are correctly accommodated for.^5^ However, current methods are often highly susceptible to intrinsic motion artifacts and visual noise is common due to the nature of clinical B-mode recordings^6,7^ This often limits accurate measurements to formal laboratory environments, where both the individual’s limb and ultrasound probe are constrained in customized equipment to limit motion.^8,9^

During FMD measurements, baseline artery diameter is first recorded. Downstream blood flow is then occluded for 3-5 minutes with a blood pressure cuff.^10^ Upon release of occlusion, increased endothelial shear stress induces vasodilation. The degree of dilation can be measured by monitoring peak post-occlusion arterial diameter, with percent FMD serving as a reliable biomarker for endothelial health. Able to be completed in approximately 10 minutes, this estimate of vascular health has proven useful to stratify risk of clinical cardiac events^11^ and is commonly employed in the research setting.^12^

Past software specifically for FMD measurements rely heavily on user input to define a region of interest and center point in the artery.^6,7^ If these user-defined parameters are inconsistent or if the vessel is recorded at an angle, the associated Canny edge detection within the program is unable to accurately measure vessel diameter.^13^ This can lead to potential loss of correctly read frames or inaccurate estimates of vessel diameter (Figure 1). This may introduce minimal potential error and frame loss at baseline when vessel movement is limited. However, post-occlusion measurements in FMD have intrinsically more movement, making this key recording parameter potentially fraught with inaccuracy. As the difference from 5% to 15% change in FMD equates to either significant pathology or a normal response, accuracy and reproducibility of these measurements is key.^5^

**Figure 1:**
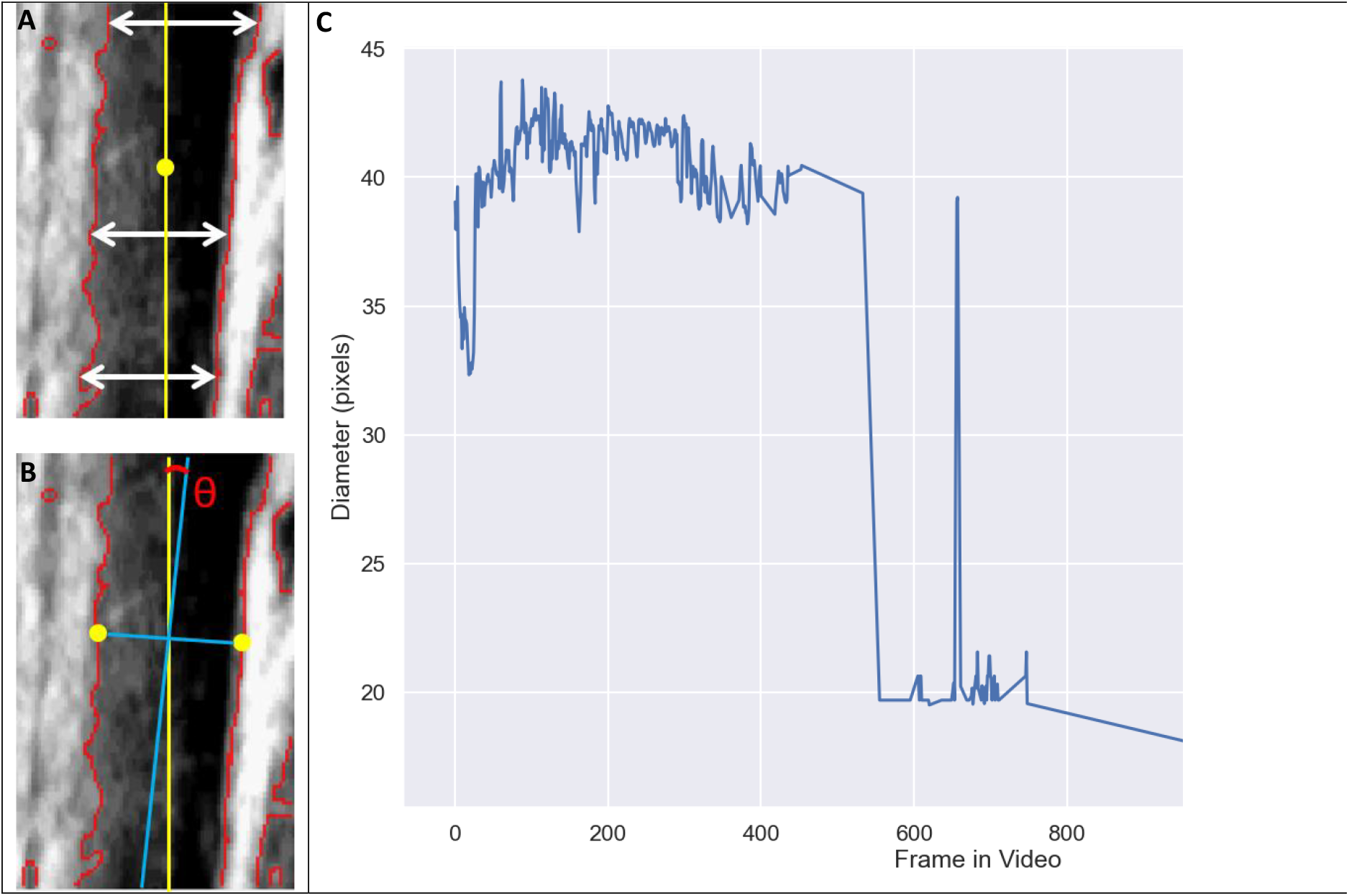
**A/B)** Summary of how FloWave.US detects vessel wall edges (red outline), fits an angle (Ө), and measures row by row diameters to average. **C)** Example of inaccuracies in dynamic vessel measurements noted from clinical ultrasound due to frame dropout when using this technique.

Herein, we introduce BWave.US, a new open-source program written in Python for post clinical recording analysis. This offers an alternative for accurate dynamic blood vessel diameter measurements which is deployable with clinical resources. A key improvement over past programs is the reduced number of steps requiring user-input to maintain reproducible results. Further, this program employs a similar open-source approach to facilitate easy integration with other coding languages and software, while also being compatible with shear rate approximations^14,15^ and ability to perform allometric scaling.^16^ We hypothesized that BWave.US preserves more usable trials (eclipsing a 70% frame capture threshold) than the well accepted, common research standard, FloWave.US, and is less variable in diameter measurements attained between independent raters for clinically gathered ultrasound recordings.

## Methods

Data for software refinement was attained from a previous study of 38 individuals aimed at understanding basic vascular physiology, and thus not registered on clinicaltrials.gov. This study was approved by our Institutional Review Board at Spaulding Rehabilitation Hospital and all participants completed informed consent. Participants were either healthy uninjured controls or had a traumatic spinal cord injury at least 1 year prior, were ages 18-50, had their injuries classified using the American Spinal Injury Association Impairment Scale (AIS)^17^ as A-D, and with neurological level of injury at or above T6. We chose these diverse cohorts to identify how BWave.US performed under typical study conditions with a range of expected vascular response (from normal in controls to a spectrum of pathology in those with spinal cord injury) and compare our results to a popular, laboratory-based software for FMD measurements, FloWave.US.

B-mode ultrasound recordings (Philips iu22 ultrasound imaging system, Bothel, WA) were taken at the brachial artery at a point 5 cm proximal to the antecubital fossa, and at an insonation angle of < 60°, over a recording of 10 seconds to measure mean baseline diameter. Shear rate was calculated as the 10 second average of peak flow divided by mean diameter. Recordings were performed by an experienced clinical ultrasonographer, though neither the limb, nor the ultrasound probe were constrained. After baseline measures of brachial artery diameter, a forearm cuff was placed 1 cm distal to the antecubital fossa and inflated to vascular occlusion for 5 minutes to assess subsequent endothelial health through FMD. Post-occlusion, shear rate was similarly measured over the 5 maximal beats of dilation. B-mode recordings were again collected for three minutes immediately following this to record the post-occlusion brachial artery diameter. These dynamic videos were selected to assess software refinements in a clinically representational and challenging environment.

After identifying the current arterial wall detection shortcomings with FloWave.US (Figure 1), open-source BWave.US was created (https://github.com/bishyboi/BWave). BWave.US detects vessel walls for diameter measurements from clinical vascular ultrasound recordings. It accomplishes this by first preprocessing frames with equalized contrast and Gaussian blur^18^ using OpenCV Sobel edge detection^19,20^ and Otsu Binary thresholding.^21^ This, thereby informs Watershed image segmentation, performed in sci-kit image.^22,23^ Contacting pixels are grouped contiguously, and two vessel edges are identified from the straightest pixel groups, determined using the sum of residuals for each pixel group (1)(Figure 2). Distance between vessel edges are calculated by fitting two parallel lines and calculating the distance between them (2).

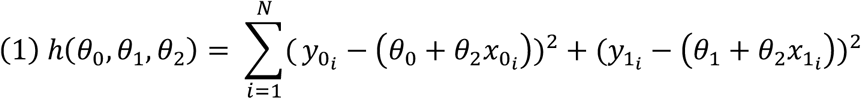

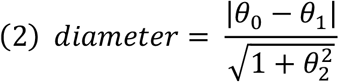

**Figure 2:**
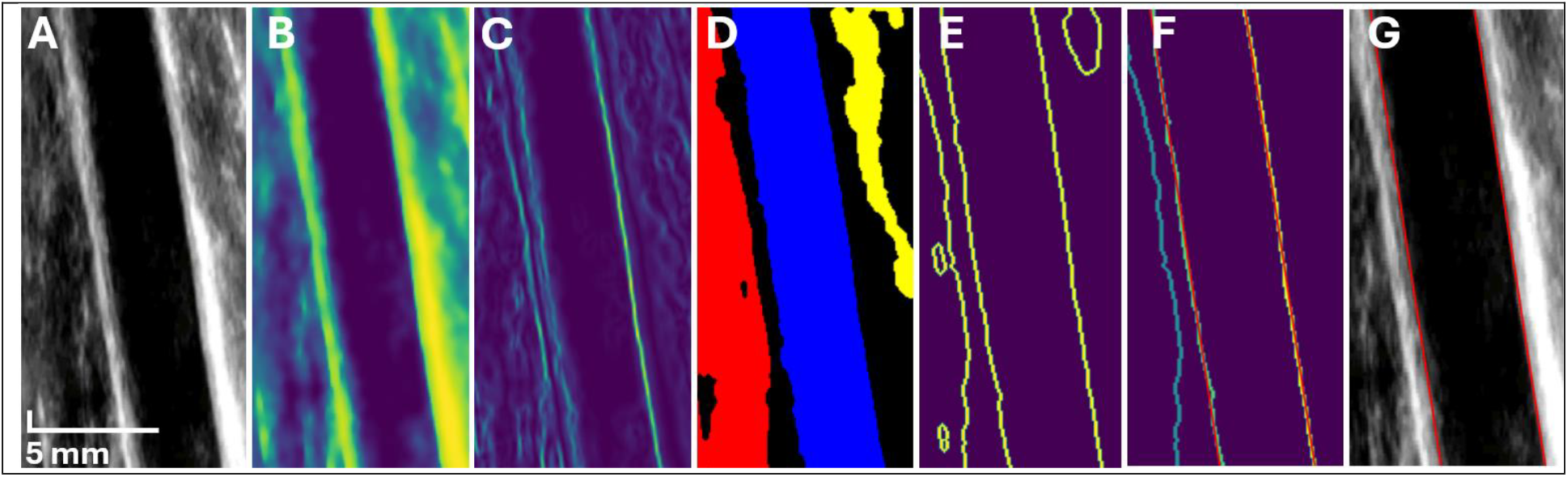
Ultrasound image (**A**) preprocessing with equalized contrast and a Gaussian blur (**B**). Sobel edge detection is applied (**C**) to inform image segmentation. Otsu’s binary thresholding and Watershed image segmentation is applied to separate discrete parts of the image (**D**). Grouped pixels are then identified (**E**) and cleaned with 75% image height threshold based on the assumption that the imaged vessel should span the height of the frame. Pixel groups are identified, and the two linear groupings (vessel walls) are fit with a residual sum of squares line to best approximate vessel walls for accurate diameter measurements (**F/G**).

After analyzing each frame, signals are filtered with a second-order Butterworth digital high-pass filter with a cutoff frequency of 0.05 Hz applied using the Python library, *SciPy*,^24^ to eliminate low-frequency noise and drift in the physiological data. Next, data points exceeding 0.9 standard deviations from zero were removed to reduce large motion artifacts. A second round of outlier removal was then performed on the remaining data, excluding any points that deviated more than two standard deviations from the mean of the filtered signal. The region of interest data is saved into the file with the raw, unfiltered diameter. This ensures that results can be duplicated and allows future users to make their own decisions with data filtering.

Accurate vessel wall detection facilitates these diameter measurements, wherein FMD can be calculated. These may be reported in raw form or are available for further allometric scaling.^16^ The latter is facilitated through obtaining the slope of the log scaled baseline and post-occlusion. The allometrically scaled FMD is then calculated by the following formula (3):

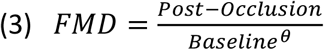

where θ represents the slope of the log-scaled baseline vs. post-occlusion. This internal check allows confirmation of key assumptions of this technique. Changes in shear rate from baseline to post-occlusion peak are further quantifiable for FMD adjustments if desired using traditional software.^6^

Comparisons between the current laboratory gold standard (FloWave.US in MATLAB) and our updated code (BWave.US) were done by modifying the FloWave.US BMode.m script to use the same region of interest for each video during head-to-head comparison. Objective comparisons were made between programs regarding video frame capture rate, reproducibility, and diameter measurements. Specifically, a 70% threshold for frame rate capture was assessed as a surrogate of a usable recording. To assess reproducibility, diameter measurements were independently performed by two team members. Interclass correlations (ICC) and root mean square error (RMSE) were calculated for baseline and post-occlusion measurements with both programs. A Bland-Altman plot^25^ was generated to assess for measurement biases between programs. Following assessments of normality, Mann-Whitney U or paired t-tests compared metrics between programs. Descriptive statistics are presented as mean ± standard deviation or interquartile range (IQR). A p-value of < was treated as significant.

## Results

We analyzed 68 total videos with both FloWave.US and BWave.US to quantify the performance of each using clinically gathered ultrasound recordings. In the control cohort, calculated FMD was 9.2 ± 8.6% using FloWave.US vs 16.4 ± 7.6% using BWave.US (p=0.08). In the spinal cord injured cohort, calculated FMD was 10.0 ± 10.9% using FloWave.US vs 6.3 ± 5.7% using BWave.US (p=0.35). Notably, these discrepancies between software results may be traced to fewer complete trial datasets available when using FloWave.US, given differing rates of achieving the specified frame read threshold. Per resting baseline recordings, FloWave.US was able to read a median of 100 (IQR, 98-100) % of all frames, whereas BWave.US read 89 (IQR, 83-94) % (p=0.0001). In the post-occlusion setting, FloWave.US read 93 (IQR, 66-100) % of all frames, whereas BWave.US read 86 (IQR, 80-90) % of all frames (p=0.91, Figure 3).

**Figure 3:**
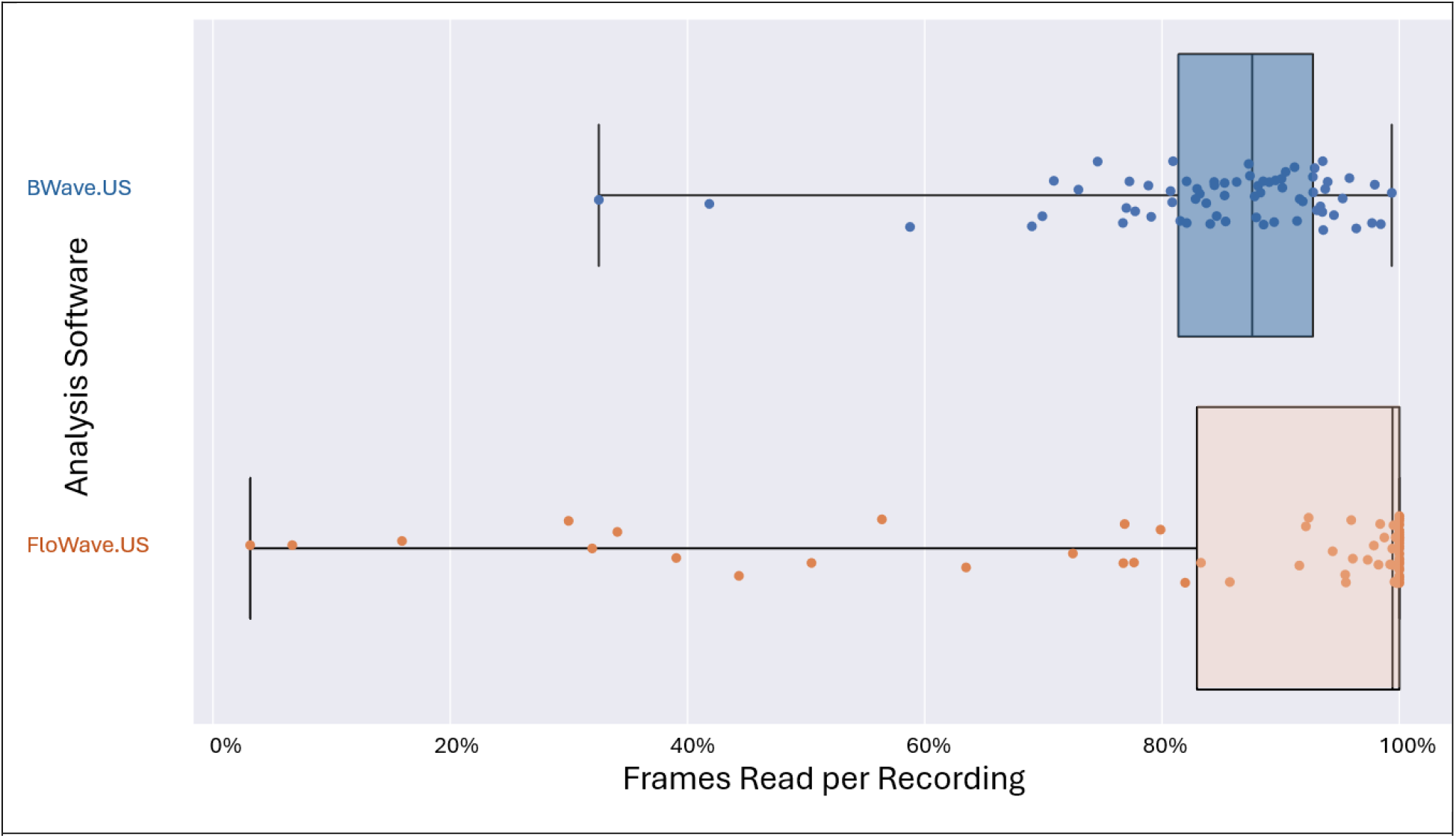
Frame capture rate for post-occlusion trials for both FloWave.US and BWave.US. 70% video frame capture rate represented by vertical dashed line.

In assessing the 70% frame read threshold for usability for each program, both FloWave.US and BWave.US surpassed this in 97% of baseline trials. For post-occlusion measurements, FloWave.US met the frame read threshold in 76% of videos compared to 97% for BWave.US (p=0.01, Figure 3). As calculations of FMD require both a usable baseline and post-occlusion measurement, this practically translated to 9 complete trials lost for 34 participants when employing FloWave.US and 2 complete trials lost when utilizing BWave.US (p=0.02).

In assessing interrater reliability, baseline measurements using FloWave.US demonstrated moderate consistency between raters (ICC=0.51, RMSE=0.76 mm). Reliability was greater using Bwave.US (ICC 0.82, RMSE=0.38 mm), with significantly less RMSE (p=0.01). In post-occlusion interrater reliability measurements, this relationship between FloWave.US (ICC=0.69, RMSE=0.60 mm) and Bwave.US (ICC=0.77, RMSE=0.44 mm) was similar, though RMSE was not significantly different between software (p=0.20). Grouping all interrater reliability diameter measurements by software analysis method, there was no significant difference in RMSE between groups (p=0.41).

Bland-Altman analysis demonstrated that BWave.US had a fixed positive 4.43-pixel bias in diameter readings when compared to FloWave.US (R^2^=0.28, p<0.00001, Figure 4). This equates to a mean difference of 0.35 mm between analysis software.

**Figure 4:**
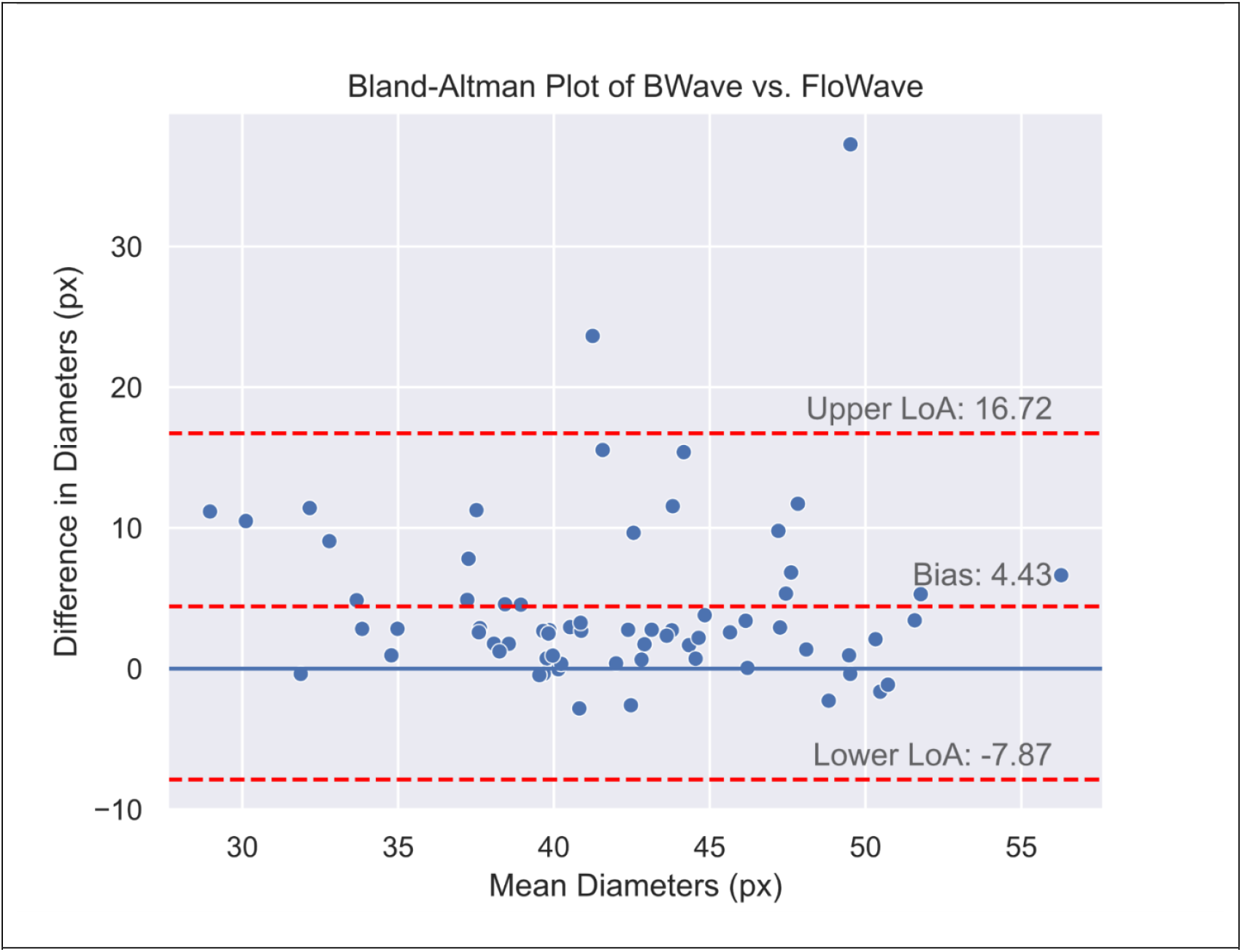
Bland-Altman Plot demonstrating fixed bias of 4.4 pixels between FloWave.US and BWave.US programs for dynamic ultrasound measurements of the brachial artery during flow mediated dilation.

## Discussion

In comparing the two programs for FMD, FloWave.US, the current laboratory gold standard, preserved more readable frames under baseline conditions where there was minimal motion artifact. There were notably more near 100% reads using this program. However, in the post-occlusion condition, which created motion artifacts due to clinically important dilation of blood vessels, BWave.US outperformed FloWave.US. This critically improved complete usable, clinically obtained dataset measurements from 74% from FloWave.US to 94% with BWave.US. This is a key improvement with BWave.US and should help facilitate important FMD measurements of vascular health to be performed more inclusively in clinical settings.

Commonly with FMD measurements there may be significant variability in edge-detection, and as such measurements are repeated up to ten times to ensure reproducibility.^1,2^ The automation of this process in BWave.US, with saved regions of interest coordinates, thus represents an important feature. This has potential to save time of measurements, aiding in translation of the FMD technique to clinical settings.^9,26^ This saved region of interest additionally enables users to replicate results consistently and removes variability from user input. BWave.US saves diameter measurements in their raw, unfiltered form, giving users the flexibility to apply their own data filtering methods or allometric scaling.

Even when region of interest is re-defined between raters, independent diameter measurements showed higher intraclass correlations using BWave.US, though lower values of RMSE were only statistically significant for baseline measurements. This improvement in reliability is likely to generate more consistent results when analyzing clinical data, though falls short of agreement seen in tightly controlled laboratory testing.^6,27^ This is, in part, due to repeated manual definition of the region of interest. Mindful of this, future iterative improvements of this BWave.US open-source code are encouraged.

Furthermore, our Bland-Altman analysis demonstrated that BWave.US had a fixed positive bias in diameter. This limitation should be considered when using this analysis software. For a typical brachial artery diameter of 4.9 mm,^28^ a normal 15% FMD would be underestimated by this fixed bias to be 14%. Reassuringly, a proportional bias was not present, and as pre/post measurements are collected and percent change calculated, a fixed bias is of less practical consequence. These Bland-Altman calculations should be interpreted in the context of the high frame rate dropout for FloWave.US influencing results, which by convention was treated as the gold standard.

### Limitations

BWave.US was developed and tested on a limited dataset of 38 individuals, and as such, there is a risk of overfitting. This potentially limits its broader clinical effectiveness, and future replication studies on differing datasets to verify the findings would be helpful. This open-source analysis software provides reproducible flow mediated dilation results and holds translational promise for improving access to measurements of vascular health using clinical equipment and expertise.

## Conclusions

FloWave.US proves to be useful when there is little-to-no expectation for change in blood vessel diameter, but BWave.US provides an improved alternative when more motion artifact is present in the video or vessel dilation is expected.

## Data Availability

All code is open source. All data produced in the present study are available upon reasonable request to the authors.

## Notes

### Competing Interest Statement

The authors have declared no competing interest.

### Funding Statement

This study was funded by the Foundation for PM&R. Dr. Solinsky's time was protected by K23HD102663
through National Institutes of Health /National
Institute of Child Health and Human Development
(NIH/NICHD).

### Author Declarations

IRB of Mass Gen Brigham gave ethical approval for this work.

